# Characterising *Streptococcus pneumoniae* Transmission Patterns in Malawi Through Genomic and Statistical Modelling

**DOI:** 10.1101/2024.11.22.24317796

**Authors:** Rory Cave, James Chirombo, Uri Obolski, Sophie Belman, Akuzike Kalizang’oma, Thandie S. Mwalukomo, Arox Kamng’ona, Comfort Brown, Jacquline Msefula, Farouck Bonomali, Roseline Nyirenda, Todd D. Swarthout, Brenda Kwambana-Adams, Neil French, Robert S. Heyderman

## Abstract

Controlling the carriage and transmission of *Streptococcus pneumoniae* in children from high-disease burden countries is crucial for disease prevention. To assess the rate of spread, and the factors associated with the high frequency of transmission despite pneumococcal conjugate vaccine (PCV) introduction, we measured evolution divergence time using the whole genome sequences of *S. pneumoniae* collected from 1,617 child participants from Blantyre, Malawi between 2015 and 2019. These children included both PCV13 vaccinated children aged 2 to 7 years and PCV13 unvaccinated children aged 5 to 10 years who were age ineligible when PCV was introduced. Using a generalized additive mixed model (GAMM) and relative risk (RR) frameworks, while accounting for household geospatial distances, we found that the spread of lineages became widespread across the population of Blantyre over approximately four years, with transmission being more likely between neighbouring households. Logistic regression and random forest models predicted a higher incidence of events among preschool children in densely populated, higher socioeconomic areas. Additionally, recent transmission was linked to recently expanding, non-vaccine serotype lineages that are penicillin non-susceptible. Our findings suggest that enhancing vaccine-mediated immunity among preschool-aged children in high density settings could reduce transmission of disease-causing and antimicrobial-resistant pneumococcal lineages, therefore strengthening herd protection for vulnerable individuals (e.g. very young children and people living with HIV).

## Introduction

Understanding the transmission dynamics of respiratory microbes is crucial for effectively targeting public health interventions aimed at controlling person-to-person spread. Mathematical models integrating human, environmental, and pathogen-related characteristics have been widely used to study the spread of SARS-CoV-2 and *Mycobacterium tuberculosis* in both local and global contexts (1–4). However, transmission modelling becomes increasingly complex when investigating a diverse bacterial species such as *S. pneumoniae* which consists of multiple co-circulating within the same environment.

*Streptococcus pneumoniae* (the pneumococcus) is a respiratory pathogen responsible for a high global burden of pneumonia, meningitis, and sepsis, associated with approximately 300,000 deaths annually among children under five years (5,6). Pneumococcal nasopharyngeal carriage is typically asymptomatic, but is a prerequisite for both transmission and disease (7). Transmission occurs through direct person-to-person contact via respiratory droplets, particularly amongst children and in crowded settings (8–10). There are over 100 serotypes, and 900 lineages defined by their Global Pneumococcal Sequence Type (GPSC). These strains frequently co-circulate within a single region, with multiple strains often carried simultaneously in the human nasopharynx, particularly in resource-limited settings (11,12).

Pneumococcal conjugate vaccines (PCV) has been introduced into the routine immunisation programme of over 160 countries, reducing pneumococcal carriage, transmission, and disease (7,13). However, despite robust direct protection, control of person-to-person spread by PCVs and therefore herd immunity has been incomplete in many settings. Mathematical models in conjunction with pneumococcal genomic data have been used to determine the rate of spread across countries, transmission rates from mother to child, and the impact of vaccination on incidence of invasive pneumococcal disease (IPD) across different age groups in different settings (14–18). While the factors associated with pneumococcal carriage have been extensively studied (19–21), less is known about the epidemiological and bacterial mechanisms of spread within densely populated urban areas following PCV introduction.

We have previously shown that in Malawi, following the routine introduction of PCV13 in 2011, there has been limited herd protection against IPD, particularly for unvaccinated children and adult persons living with HIV (PLHIV) (22–25). Despite PCV coverage exceeding 90%, this limited impact may be attributed to the persistence of high pneumococcal vaccine serotype carriage in the population (24), which - along with age-related factors - contributes to a sustained high force of infection (26). Furthermore, shifts in the pneumococcal population structure have led to the emergence of genotypes with virulence and AMR profiles that confer competitive advantage, as well as pneumococcal capsule locus variant lineages that retain their serotype (PCV13 serotypes 3, 14, 23F) and contribute to vaccine escape (27,28). We hypothesise that the emergence and persistence of vaccine-escape lineages are driven by short-range transmission among young children, amplified by antimicrobial resistance (AMR) related to a high rate of antibiotic exposure (29).

To test this hypothesis, we integrated large-scale longitudinal genomic and epidemiological data from a high burden urban population in Malawi. By analysing divergence times, and geographical locations of pneumococcal genome pairs and employing machine learning and statistical models, we infer the time required for pneumococcal lineages to reach saturation and become fully mixed within the community. We evaluate the likelihood of transmission between neighbouring and distant households, and identify key factors associated with transmission, including child age, population density, vaccine serotype, penicillin non-susceptibility, and GPSC lineage.

## Results

### Modelling the time *S. pneumoniae* spread to reach saturation, mixing points, and the likelihood of transmission between neighbouring and distant households

To explore pneumococcal rate of spread within the population and determine which human and bacterial factors are associated with transmission in urban Blantyre, Malawi, we have used the Pneumococcal Carriage in Vulnerable Populations in Africa (PCVPA) dataset, collected from 2015 to 2019 (Table S1 and Figure 1) (24). This dataset consists of 2,283 child participants in which a single isolate from each was participants sequenced, comprising 59 unique serotypes, of which 23.1% (n=528) are PCV13 VT. There are 118 GPSC lineages, with 37.4% (n=854) of isolates being non-susceptible to penicillin defined by a minimum inhibitory concentration (MIC) > 0.12 μg/ml for meningitis infections), 30.3% (n=692) resistant to tetracycline, and 16% (n=366) resistant to erythromycin.

**Figure 1:**
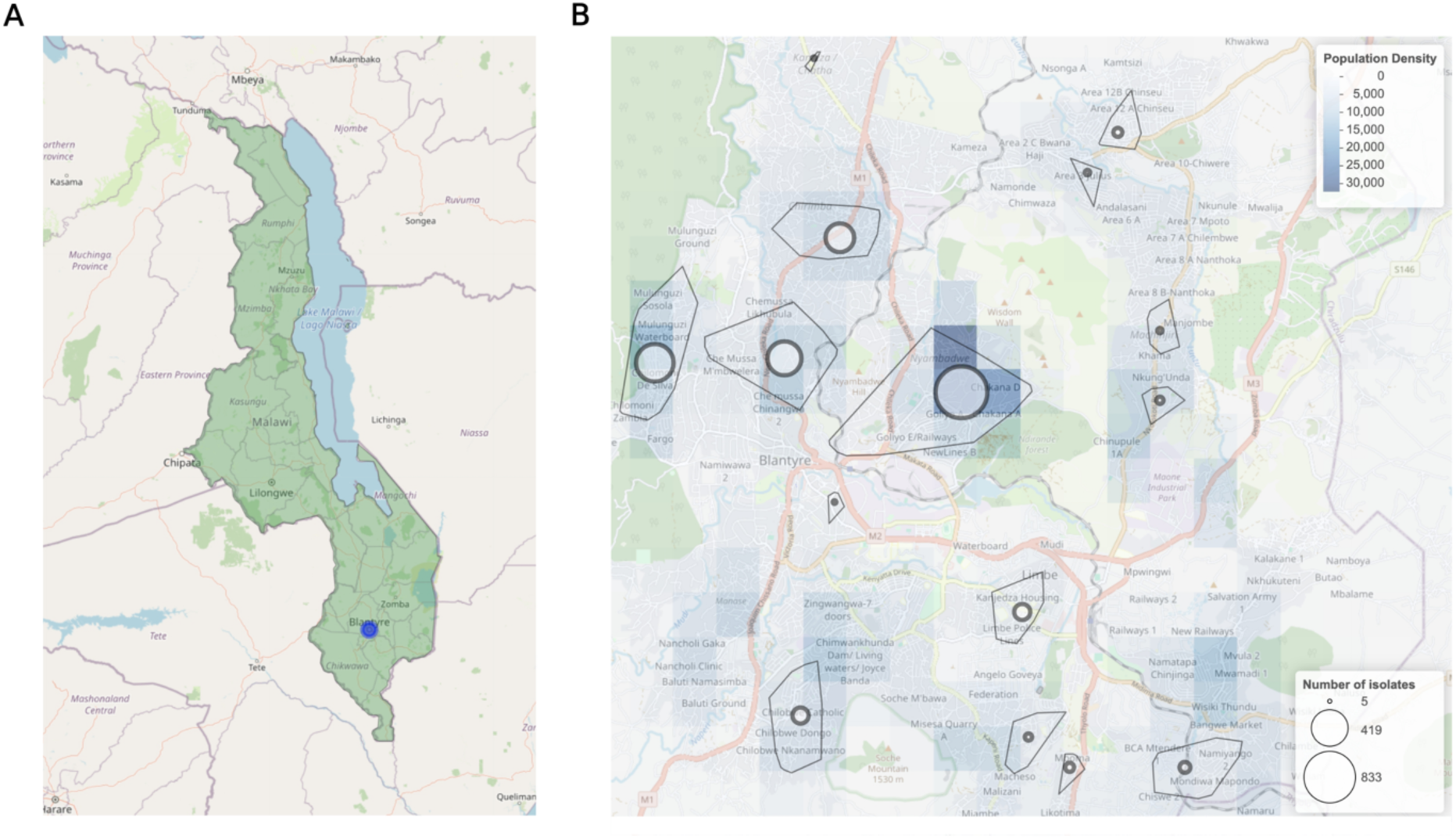
Geographic representation of Malawi and Blantyre. **A)** Map of Malawi indicating the location of Blantyre (Blue dot). **B)** Detailed map of Blantyre illustrating population density, with areas where samples were collected outlined by black convex hulls, and the number of isolates collected from each area represented by marker size.

To determine the pairwise divergence times between S. *pneumoniae* carriage isolates among children in the community, we first generated a Bayesian time-calibrated phylogenetic tree using BactDating for each GPSC lineage with recombination removed. We successfully constructed a Bayesian time-calibrated phylogenetic trees for 31 out of the 118 GPSCs. These lineages comprised 1,617 of the 2,283 carriage isolates collected from the PCVPA dataset, which showed no geographical clustering of lineages (Table S1).

Using pairwise divergence times between isolates derived from the Bayesian time-calibrated phylogenetic tree, we developed a generalized additive mixed model (GAMM) to assess the relationship between divergence time and distance between pairs (Figure 2A). The model explained 76.6% of the variance with a statistically significant relationship between the divergence time and geographical distance (p < 0.001). Initial the GAMM model predicts an increase in divergence time as geographical distance between pairs increased. This confirms the convention that pneumococcal spread among children is relatively localised, only gradually spreading to the rest of the community over time. Notably, the trend reached saturation at 3.92 years of divergence, with pairs being 2.31 km apart on average (the mean distance of all pairs within 10 years of divergence was 2.46 km, ranging from 0 -14.3 km). These findings suggest that even in a high density, high carriage prevalence population, pneumococcal community spread occurs relatively slowly.

**Figure 2:**
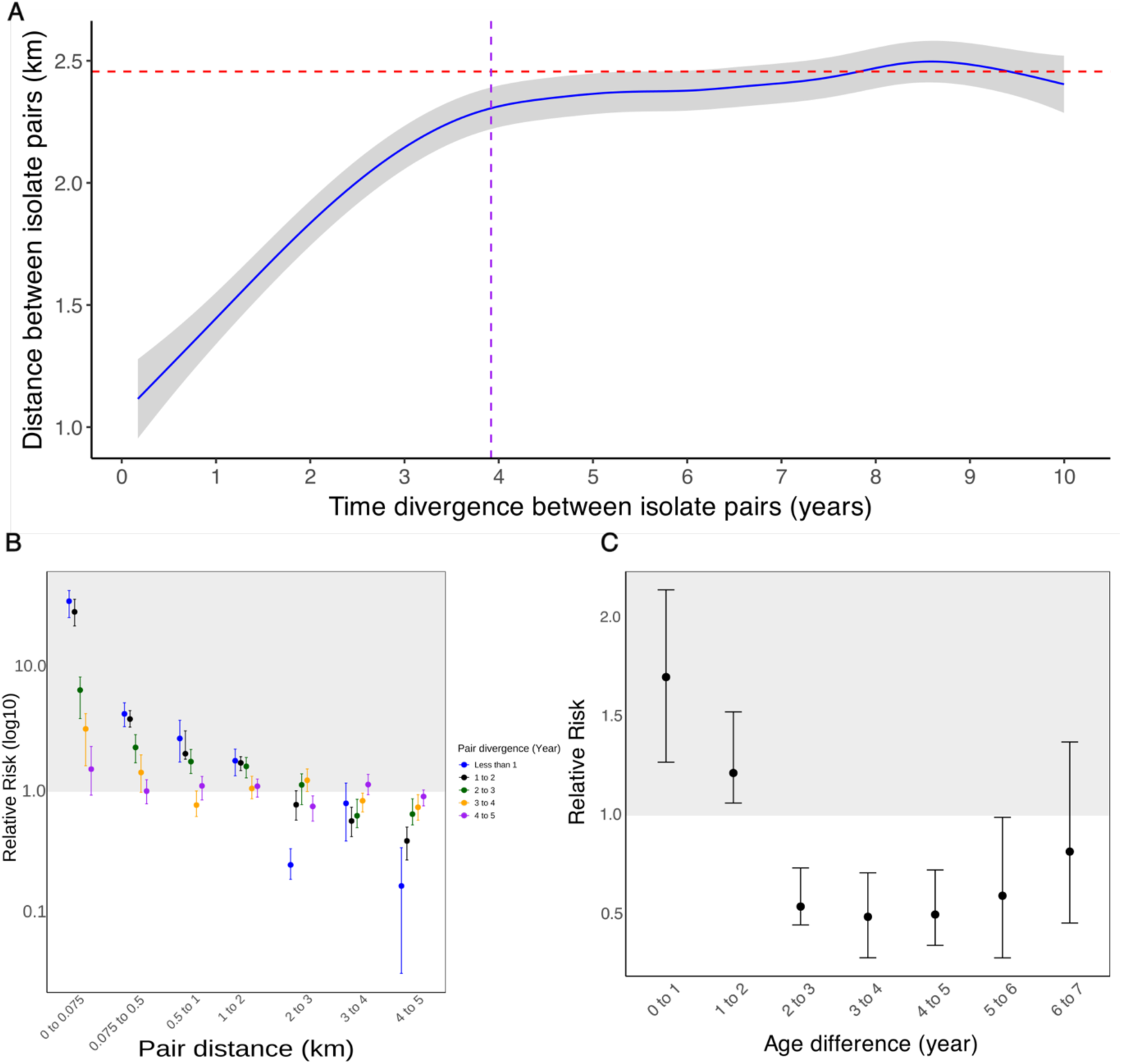
GAMS model and RR analysis. **A)** A GAM mixed model showing the divergent time between pairs against the distance between pairs. Blue line is the plotted GAM model, grey area is the 95% confidence interval, purple dashed line is the saturation point of the curve and red dashed line is the mean distance of all pairs that have less than 10 years of divergence. **B)** Relative risk of isolates pairwise divergence times difference can be found between different pairwise distance. **C)** Relative risk of isolates pairwise divergence of less than 1 year divergence found between pairwise difference of children ages.

To further characterise the transmission dynamics and identify potential targets for intervention, we calculated the relative risk (RR) across five divergence time groups (less than 1 year, 2 to 3 years, 3 to 4 years, and 4 to 5 years) over a range of distances (Figure 2B). We set the lowest distance to measure relative risk at 75m apart, based on the mean nearest neighbour distance of 72m, which yielded the highest RR for each divergence time group compared to other distances. With each increase in divergence time group, we observed a general decrease in the relative risk of transmission occurring at those distances, as well as a decrease in relative risk with increasing distance between pairs up to 1-2 km apart. This suggests that transmission is most likely to occur between neighbouring households, only gradually spreading across the community over time. Moreover, we observe that pairs less than 75 metres apart reached a non-significant difference from 1 (RR 1.53, 95% CI 0.92–2.34). This indicates that it takes 4 years for isolates to become fully mixed in the community in Blantyre.

We also found that the likelihood of transmission between children of different ages decreased as the age difference increased (Figure 2C). The highest RR was seen when the age difference was between 0 and 1 year (RR 1.7, 95% CI 1.27-2.2), compared to 1 to 2 years difference (RR 1.22, 95% CI 1.04-1.42) and 2 to 3 years difference (RR 0.54, 95% CI 0.35-0.84). This suggests that transmission between children is more likely to occur among children of similar ages.

### Pneumococcal transmission is decreased with age and vaccine serotype isolates, but increased with higher population density, socioeconomic score, and penicillin MIC

To further understand the role of human and bacterial factors in pneumococcal transmission within this urban community, we conducted univariate and multivariable mixed-effect logistic regression analyses, informed by the findings from the GAMM model with isolates to be recently transmitted if they were less than 4 years divergence apart and 2.5km apart (Table S2). Consistent with previous studies (30,31), the mixed effect logistic regression revealed a significant decrease in pneumococcal transmission with increasing age (adjusted OR 0.82 per year, 95% CI 0.73-0.92; p < 0.001), and a significant increase in transmission with population density (adjusted OR 1.31 per km^2^, 95% CI 1.18-1.46, p < 0.001) (table 1).

**Table 1:** Univariate and multivariate mixed effect logistic regression model on human and bacteria characteristics associated with transmission

| Characteristic | Univariate |  |  | Multivariate |  |  |
| --- | --- | --- | --- | --- | --- | --- |
|  | OR<sup>1</sup> | 95% CI<sup>1</sup> | p-value | OR<sup>1</sup> | 95% CI<sup>1</sup> | p-value |
| Age | 0.77 | 0.70, 0.86 | <0.001 | 0.82 | 0.73, 0.92 | <0.001 |
| Population density (per km<sup>2</sup>) | 1.37 | 1.23, 1.52 | <0.001 | 1.31 | 1.18, 1.46 | <0.001 |
| Socioeconomic score | 1.09 | 0.99, 1.21 | 0.092 | 1.15 | 1.03, 1.28 | 0.014 |
| PCV13 vaccine type serotypes | 0.59 | 0.47, 0.75 | <0.001 | 0.54 | 0.35, 0.83 | 0.005 |
| Penicillin MIC (µg/mL) | 1.24 | 1.04, 1.47 | 0.014 | 1.30 | 1.09, 1.55 | 0.004 |
| <sup>1</sup> OR = Odds Ratio, CI = Confidence Interval |  |  |  |  |  |  |

In relation to bacterial factors, we found significantly less transmission of pneumococcal vaccine serotypes compared to non-vaccine serotypes (adjusted OR 0.54, 95% CI 0.35-0.83; p = 0.005). Additionally, higher penicillin MIC values were positively associated with recent transmission events (adjusted OR 1.3 per μg/ml, 95% CI 1.09-1.55; p = 0.004). Additionally, we observed a significant increase in penicillin MIC from those collected between July 2017 to June 2019 compared to those collected between July 2015 to June 2017 (Wilcoxon, p <0.001) (Figure S1). These data support the emerging evidence (18), that in a population with high vaccine uptake and high antimicrobial usage, pneumococcal lineages that are able to escape vaccine-induced immunity and that exhibit AMR are likely to spread within a community (32) .

Overall, we observed the expected pattern of higher carriage prevalence among children from the lowest socioeconomic households (table 2). However, our analysis of the impact of socioeconomic status on recent transmission revealed an unanticipated aspect of this complex process: transmission is more likely to occur among children from higher socioeconomic households (adjusted OR 1.15 per socioeconomic households score, 95% CI 1.03, 1.28; p = 0.014).

**Table 2:**
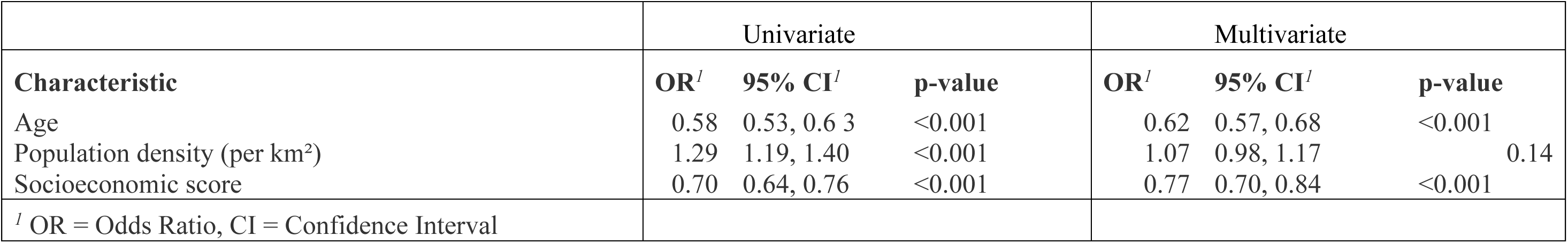
Univariant and multivariant Logistic regression model on human factors associated with carriage

### Higher transmission rates among preschool children and the identification of key GPSC lineages associated with increased transmission

Many compartmental models of infectious disease dynamics assume that transmission rates are directly proportional to the densities of susceptible and infected populations (33). However, after vaccines are introduced, there is a decline in the number of individuals that are susceptible to infection, leading to saturation. The non-linear dynamics of pathobionts such as *S. pneumoniae* are complex and somewhat unpredictable (10). To address these complexities, we developed a random forest model to identify the non-linear patterns associated with community transmission. The random forest model achieved ROC-AUC score 0.70 (CI 0.64-0.76) on an independent test data (Figure 3A). The precision-recall AUC or the test data was 0.69 (CI 0.62- 0.76) (Figure 3B). This resulted in sensitivity score of 0.69 (CI 0.61-0.76), specificity 0.55 (CI 0.48-0.63) and a G-mean of 0.62 (CI 0.57-0.67) for the test data. These metrics suggest that the model performs reasonably well, balancing sensitivity and specificity while minimising overfitting.

**Figure 3:**
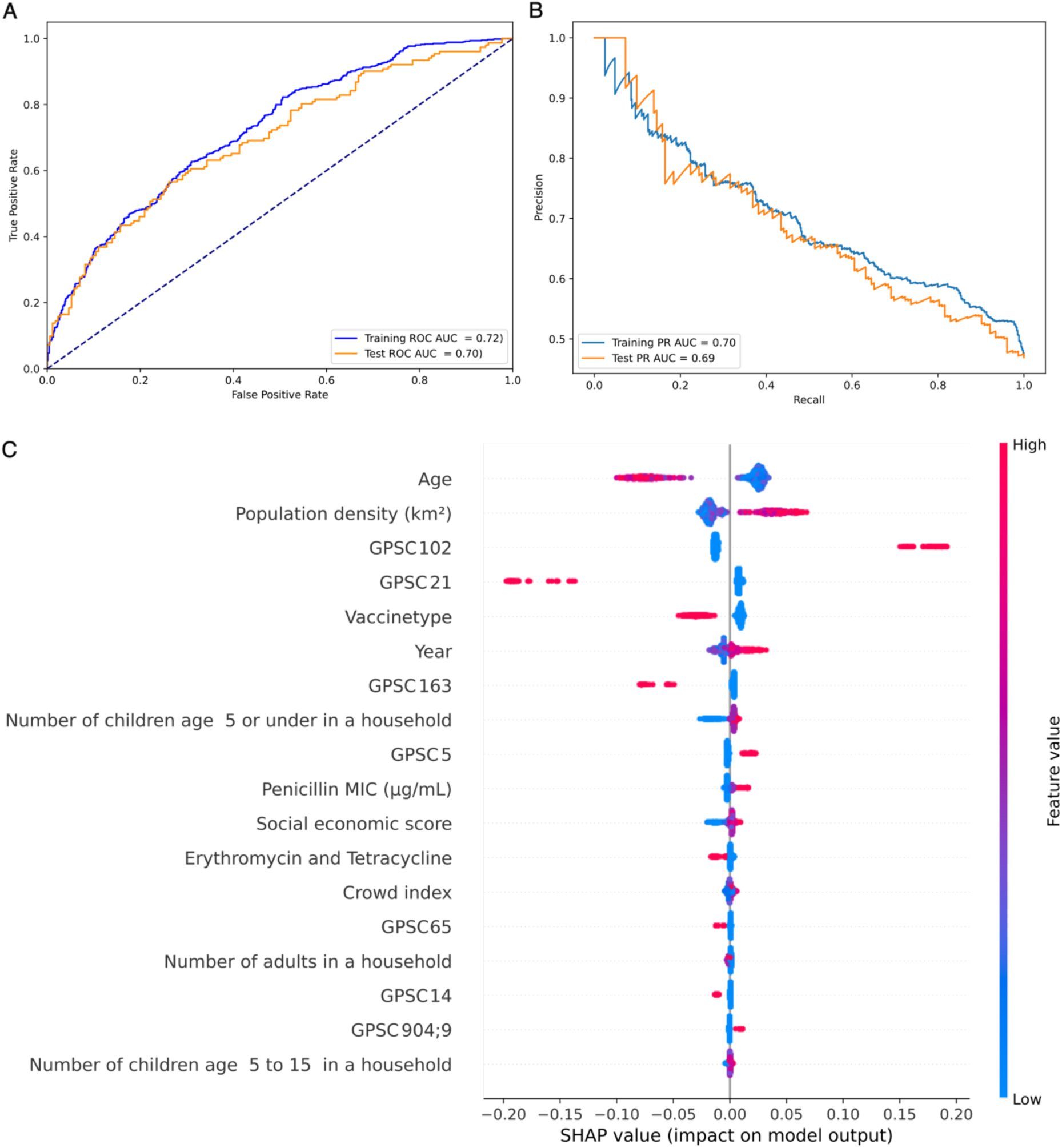
Random forest model fit to predict human and bacterial factors associated with transmission. **A)** The Receiver Operating Characteristic (ROC) curve showing the performance of the random forest model against the training and test datasets. **B)** Precision-recall curve of the random forest model on the training and test datasets. **C)** Beeswarm plot of SHAP values for each feature’s impact on the model’s predictions regarding the likelihood of an isolate being part of recent transmission. The features are displayed in descending order of importance from top to bottom, based on the average absolute SHAP value.

Using the SHapley Additive exPlanations (SHAP) values which indicate the directionality of influence of the variables on the model prediction, we identified the top five important features from the random forest model (Figure 3C). These features include both human and environmental factors such as the age of the child and population density, as well as three bacterial factors - lineages GPSC102, GPSC21, and vaccine serotype. This suggests that a combination of human and bacterial factors plays a crucial role in pneumococcal transmission within the community.

In the random forest models, we applied the same predictors used in the logistic regression analysis. Although the trends were consistent with the logistic regression, they were not strictly linear. For example, the partial dependence plot for age shows that the model identified that children aged younger than six have substantially more positive influence the model’s prediction of transmission, relative to older children (Figure 4A). In Malawi, children six years and older typically attend school, highlighting that pneumococcal transmission primarily occurs among younger, preschool children within this community.

**Figure 4:**
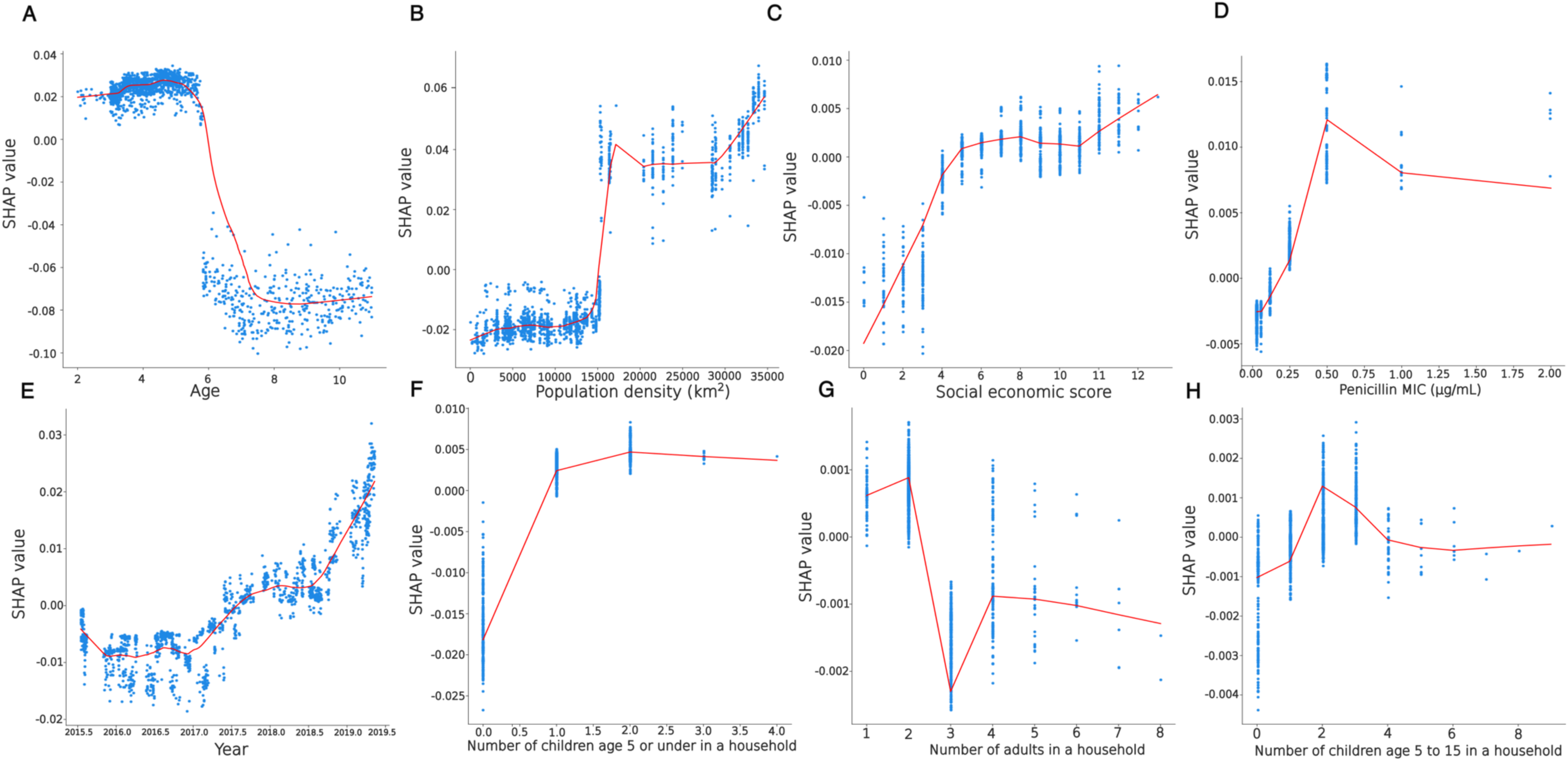
Partial plot of SHAP values for **A)** Age of child, **B)** Population density, **C)** Socioeconomic score, **D)** Penicillin MIC, **E)** Year of isolation, **F)** Number of children aged 5 or under in a household, **G)** Number of adults in a household, **H)** Number of children aged 5 to 15 in a household. The red line is the locally estimated scatterplot smoothing (LOESS) trend of the partial plot.

Regarding population density, our model predicts a slight increase in transmission around 15,000 people/km², followed by sharper rise and then a plateau between 15,000 to 30,000 people/km². Transmission increases again over 30,000 people/km² (Figure 4B). This non-linear trend suggests higher contact rates among children in higher-density populated areas contribute to pneumococcal transmission. Other factors, such as community infrastructure, behaviour and the age of children in these areas may also play a role.

Like the logistic regression model, the random forest model predicts that transmission increases with higher socioeconomic background but in a non-linear manner. Rates level off between a social score of 6 and 11 before increasing above 12, indicating possible factors associated with socioeconomic background (Figure 4C). However, as before this observation may be artefactual, as the model does not account for non-carriage events.

The random forest model also predicts that transmission increases with higher penicillin MICs but drops in for isolates with an MIC of 0.5 μg/ml or more (Figure 4D). This suggests that while penicillin nonsusceptibility may confer a fitness advantage, high penicillin MICs become detrimental to isolates, potentially because mutations in the penicillin-binding protein affect cell wall synthesis (34). However, the small number of isolates in the population with an MIC higher than 0.5 could introduce noise into these predictions. In contrast, resistance to erythromycin and tetracycline did not substantially affect transmission (Figure 3C). Indeed, while penicillin nonsusceptibility increased over the surveillance period, resistance to erythromycin and tetracycline remained stable (Figure S2).

To further understand how pneumococcal transmission dynamics have evolved in this population, we included year of isolation as a variable in our model (not included in the logistic model due to an increase in the AIC score, indicating a poorer model fit). We observed an increase in transmission between 2017 and 2018, stabilising around mid-2018 before rising again post-2019 (Figure 4E). This pattern aligns with the observed significant increase in penicillin-non-susceptible strains after 2017 (Wilcox test p value <0.001) (Figure S1).

We also used a random forest model to examine the effect pneumococcal transmission among children based on the number of children aged under-5-years, the number of adults, and the number of children aged 5 to 15 in their household, which were not included in the logistic regression model (Figures 4F, G and H). The results indicate that the presence of children in either age group generally increased the probability of transmission. However, households with two or more adults saw a decrease in transmission. This suggests that while more children may lead to greater interaction and higher transmission, having multiple adults might limit these interactions and reduce transmission.

### Recent expansion of lineage associated with increased transmission

To assess whether the random forest model accurately predicted the lineage effects associated with transmission, we employed statistical analyses commonly used in genome-wide association studies (GWAS) to identify lineage effects (35). In line with the top lineages of importance from our random forest model in predicted transmission, we find there is a significant lineages effect for GPSC102 (p-value < 0.001), GPSC21 (p-value < 0.001), and GPSC163 (p-value = 0.01). However, only GPSC102 exhibited an enhanced impact on transmission in the random forest model (Figure 3C). GPSC21 and GPSC163 were predicted to reduce transmission in the random forest model. At the sequence type (ST) level, GPSC102- ST4423 (p-value = 0.001), GPSC5-ST10599 (p-value = 0.02), GPSC102-ST10880 (p-value = 0.02), and GPSC5-ST10603 (p-value = 0.03) significantly impact community transmission. These STs belong to GPSC lineages that the random forest model identified as most important for predicting transmission dynamics.

To further explore why these GPSC and STs were more or less likely to be detected in recent transmission in our analysis, we investigated their change in prevalence during the study (Figure S3A). We found no significant increase in GPSC102 (p-value 0.05), whereas GPSC21 (p-value 0.3), GPSC163 (p-value 0.4), and GPSC5 (p-value 0.8) showed no significant during the study period. However, regarding STs that showed a lineage effect, there was a significant increase in GPSC102-ST4423 (p-value < 0.001, 5.52% increase in prevalence during the study), which emerged in the population in 2016, and GPSC5-ST10599 (p-value < 0.01. 2.68% increase in prevalence) (Figure S3B). However, GPSC5-ST10603 showed no significant change (p-value = 0.3), while GPSC102-ST10880 significantly decreased in prevalence during the study (p-value = 0.002, 1.86% decrease in prevalence). This decrease may be due to GPSC102-ST10880’s susceptibility to penicillin. Additionally, the decline in GPSC102- ST10880 could be driven by the emergence of GPSC102-ST4423, which was penicillin non-susceptible. The observed increase in GPSC102-ST4423, alongside the decrease in GPSC102- ST10880, suggests possible lineage replacement.

We further explored whether the most common STs within these GPSCs have recently expanded to determine if new lineages may influence transmission in the population (Figure 4). For GPSC102-ST1080 which had a significantly increase in prevalence during the survey and was found to have expanded within the last five years of the most recent sampling date, compared to ST10880, which significantly decreased in prevalence and was shown to have expanded over ten years ago (Figures 5A, B, C, D). This was also observed within GPSC5, where ST10599 significantly increased in prevalence during the study within the last eight years, compared to ST10603, which showed no difference in prevalence and was shown to have expanded over ten years ago (Figures 6A, B, C, D). Furthermore, for GPSC21, ST347 and ST10572 expanded around 60 and 40 years before the most recent sampling date, and GPSC163 ST19568 was shown to have expanded 40 years ago (Figures 7A, B, C, D). This along with the prevalence data, suggests that recently emergent lineages within the population are more likely to be shared than those that have been established in the population for longer periods of time.

**Figure 5:**
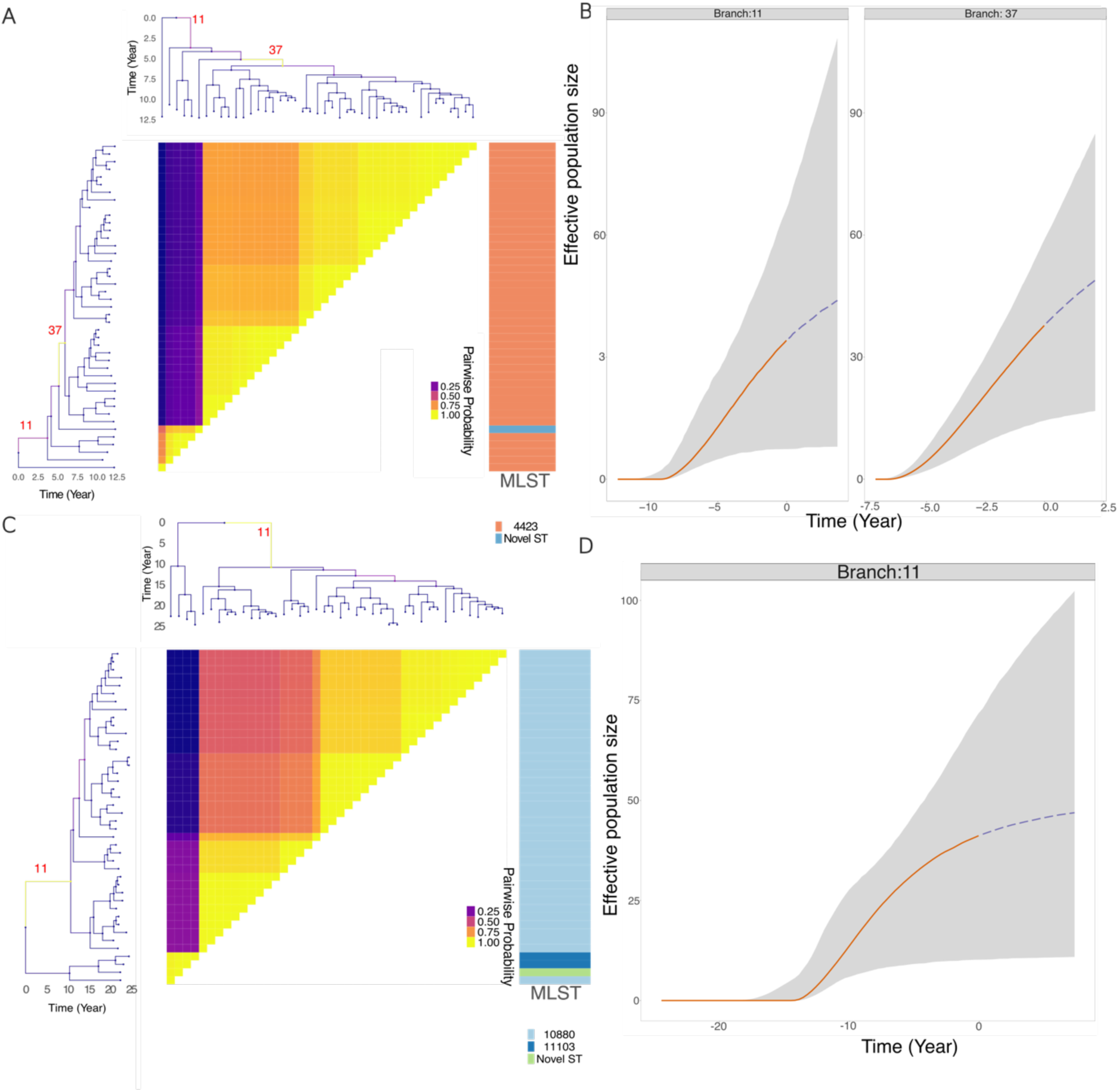
Clonal expansion and effective population of lineages showing GPSC102 lineage effects in transmission. **A)** Dated phylogeny illustrating the expansion of GPSC102-ST4423. Pairwise matrix showing the posterior probabilities of any two genomes belonging to the same subpopulation. **B)** Posterior summary of the inferred effective population size for GPSC102- ST4423. **C)** Dated phylogeny illustrating the expansion of GPSC102-ST10880. Pairwise matrix showing the posterior probabilities of any two genomes belonging to the same subpopulation. **D)** Posterior summary of the inferred effective population size for GPSC102- ST10880.

**Figure 6:**
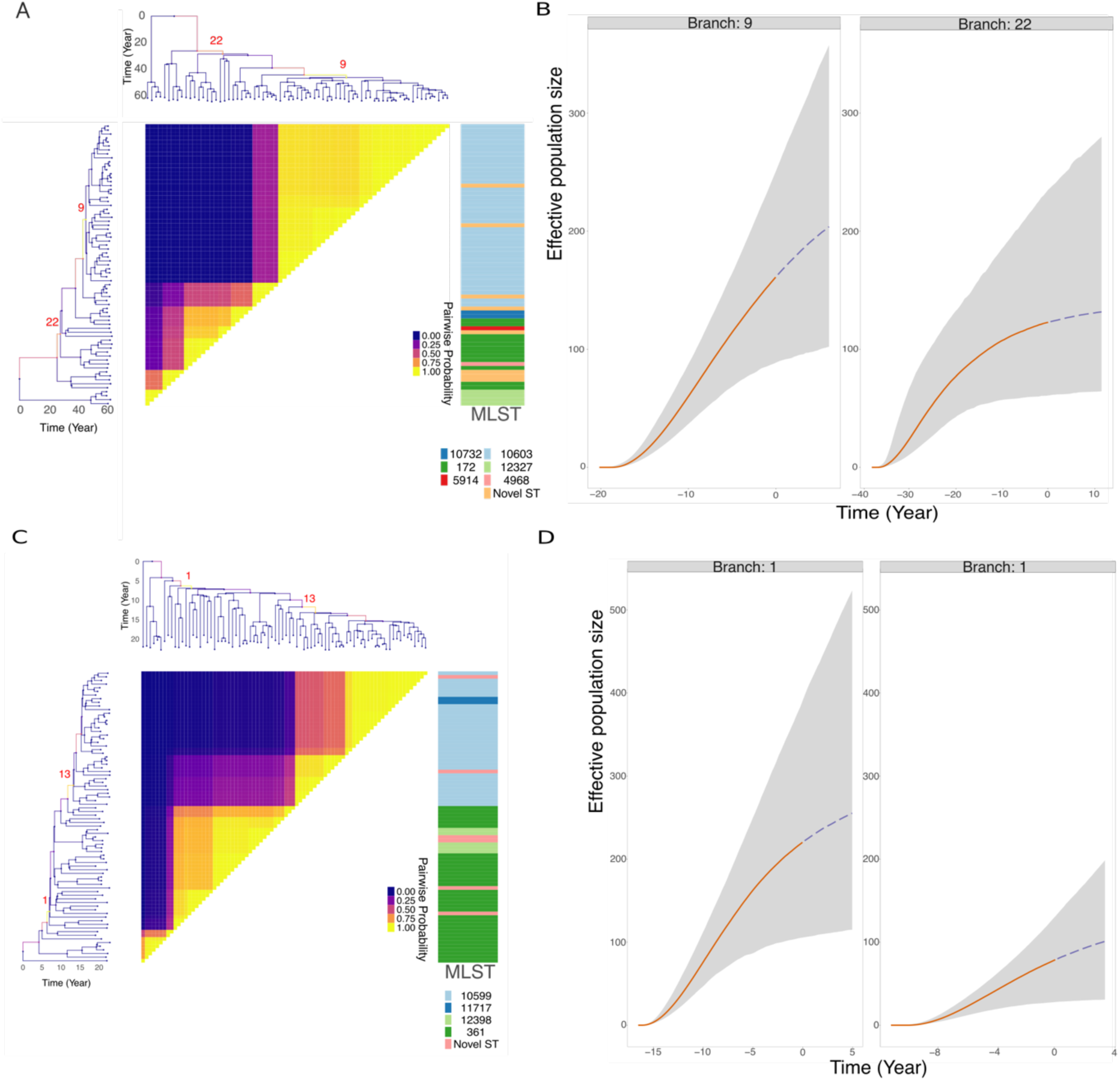
Clonal expansion and effective population of lineages showing GPSC102 lineage effects in transmission. **A)** Dated phylogeny illustrating the expansion of GPSC5-10603. Pairwise matrix showing the posterior probabilities of any two genomes belonging to the same subpopulation. **B)** Posterior summary of the inferred effective population size for GPSC5- 10603. **C)** Dated phylogeny illustrating the expansion of GPSC5-ST10599. Pairwise matrix showing the posterior probabilities of any two genomes belonging to the same subpopulation. **D)** Posterior summary of the inferred effective population size for GPSC5-ST10599.

**Figure 7:**
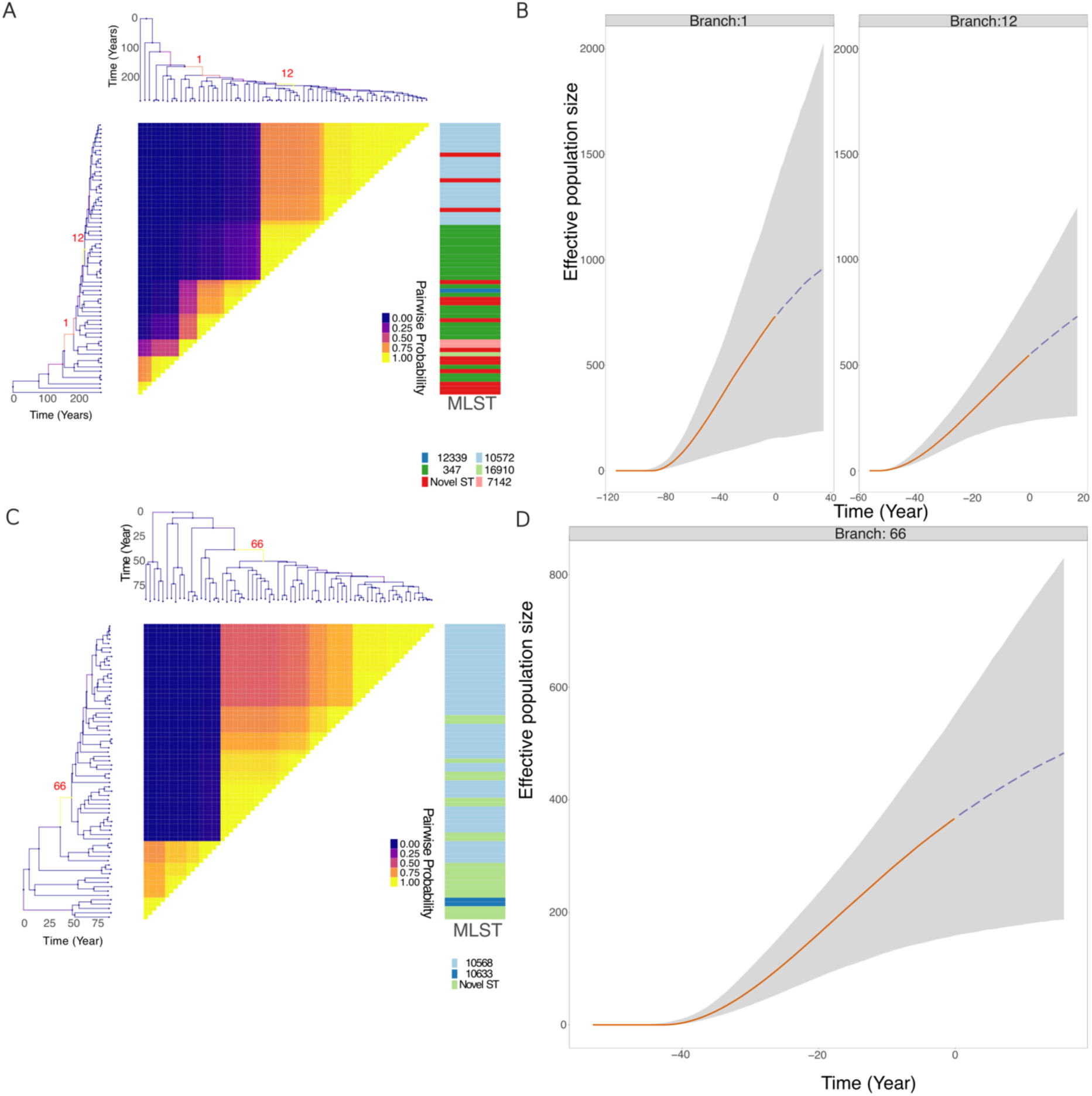
Clonal expansion and effective population size of lineages showing the effects of GPSC92 and GPSC163 in transmission. **A)** Dated phylogeny illustrating the expansion of GPSC92. Pairwise matrix showing the posterior probabilities of any two genomes belonging to the same subpopulation. **B)** Posterior summary of the inferred effective population size for GPSC92. **C)** Dated phylogeny illustrating the expansion of GPSC163. Pairwise matrix showing the posterior probabilities of any two genomes belonging to the same subpopulation. **D)** Posterior summary of the inferred effective population size for GPSC163.For the effective population size graphs, the grey area represents the 95% credible interval, and the lines denote the median. Solid lines indicate past effective population size inference, while dashed lines represent predictions of future effective population size. Point 0 on the x-axis corresponds to the most recent sample date, which was 2019

## Discussion

Using a combination of whole-genome sequencing, geospatial, and epidemiological data, our study reveals that pneumococcal transmission in Blantyre is predominantly driven by pre- school children residing in high-density areas. Transmission is largely localised, occurring primarily between neighbouring households, with pneumococcal lineages taking up to four years to fully mixed within the Blantyre community. This is driven by new emergent non-vaccine type serotypes exhibiting penicillin non-susceptibility. Our findings align with previous studies, strengthening biological relevance of our models, and provide new insights into household-to-household transmission, and the speed of pneumococcal spread in low- and middle-income cities such as Blantyre (16,21,24,36,37). Additionally, we show that emergent lineages have a greater influence on transmission dynamics compared to lineages that have been established for several years, highlighting the importance of targeted public health interventions that reduce pneumococcal transmission and so reduce invasive pneumococcal disease (IPD). Our findings also underscore the need for continuous genomic surveillance to detect potentially highly transmissible emergent lineages in such settings.

This analysis benefitted from intense sampling of 1,618 isolates over 4 years allowing us to achieve a higher resolution in tracking local transmission than in countrywide studies in Israel (1,174 isolates collected over 9 years (17)) and South Africa (6,910 isolates over 15 years (18)). Our model indicates that, even in a country with previously described high force of infection, transmission amongst children in this urban setting is relatively short range, with lineages that become fully mixed within four years (26). This is relatively slow compared to Israel, where lineages fully mixed after approximately five years, in a country with a population size of 9.6 million (population density of 434 people per km^2^ (17)). However, the slower rate of spread in Blantyre is more consistent with South Africa, with a population size 60 million (population density of 48 people per km^2^), where it took 50 years for pneumococcal lineages to fully mixed (18). This difference may reflect a variety of factors, including socio-economic status, childcare, antimicrobial usage, immune status, transport networks and local internal migration, and possibly differences in the IPD lineages or the underlying nasopharyngeal microbiome (18,21,38,39).

Data from the pre-PCV13 era suggest that infant-to-mother and infant-to-sibling transmission is the primary contributor to spread within a population (40). As might be expected for respiratory contact-dependent transmission, this was most prevalent in the higher-density areas of Blantyre, and in line with these observations, occurred most frequently between neighbouring households between preschool-aged children of similar ages. Studies in rural populations in Kenya and The Gambia, observed a high frequency of carriage episodes among young children that declined with age, likely due to increasing clearance rates (41,42). This together with the localised nature of the spread seen in Blantyre highlights the importance of close-contact interactions among young children, rather than older children and adults in driving transmission.

There are multiple bacterial factors that influence pneumococcal transmission, these include the polysaccharide capsule, which can evolve through both mutation and genetic exchange(43), is a major virulence factor and is the target for PCV. In animal models, both the type and amount of capsule has been shown to affect pneumococcal transmission dynamics (44). Following PCV introduction, new lineages have since emerged, replacing previous vaccine serotypes with non-vaccine serotypes that exhibit greater resistance to penicillin (44,45). In our study, we observed increased transmission linked to emergent, penicillin non-susceptible lineages, such as GPSC5 and GPSC102, both have expanded clonally in multiple countries following PCV introduction. In contrast, GPSC21, which contained the highest number of VTs, underwent clonal expansion before the vaccine was introduced. Together these data build on our earlier observations of persistent VT carriage (24), showing that the sub-optimal control the spread of VT lineages. Clonal expansion among a number of pathogenic bacteria with enhanced transmission capabilities has been linked to the acquisition of antimicrobial resistance (AMR) (45–48). However, the driver of the increase in prevalence and transmission of the penicillin non-susceptible pneumococcal isolates in our study remains unclear. The use of beta-lactam antibiotics has previously been linked to a rise in penicillin non-susceptible isolates, but we did not collect this data (49).

Limitations of this study include the absence of data on factors known to influence pneumococcal carriage, such as viral infections, pollution, and human contact patterns (50–52). Additionally, our model does not account for individuals without pneumococcal carriage, who may provide insight into protective human factors against bacterial transmission. Single-colony sequencing from nasopharyngeal samples may also miss important transmission links, especially in young children in Malawi who often carry multiple pneumococcal lineages (53). For example, Serotype 1, which frequently causes disease outbreaks, may be underrepresented in carriage studies when only a single colony is sequenced (16). Multi-colony metagenomic sequencing could improve our understanding of transmission, as lower-abundance resistant strains may be carried alongside susceptible strains within a single host (54).

In summary, our analysis highlights the complexity of pneumococcal transmission dynamics in Blantyre, demonstrating that both human and bacterial factors contribute to localised spread. These results highlight the need for data-driven, targeted public health interventions to reduce the incidence of invasive pneumococcal disease (IPD) by integrating epidemiological data with genomic surveillance. Future refinement of these models could be achieved by incorporating multicarriage sequence data and additional epidemiological cofactors. This would further elucidate transmission patterns and support the development of more effective vaccine strategies that target transmission of disease-causing and antimicrobial-resistant pneumococcal lineages and increase herd protection for vulnerable individuals (e.g. very young children and people living with HIV).

## Methods

### Setting and study population

The city of Blantyre (228 km^2^) is in southern Malawi with an urban population of approximately 1 million (growth rate 3.9%; overall population density 3,006 people per km^2^). Within Blantyre there are multiple high-density residential areas ranging up to 34,602 people per km^2^. Recruitment to the Pneumococcal Carriage in Vulnerable Populations in Africa (PCVPA) study was between 2015 and 2019 (PCV13 introduced into routine immunisation November 2011). PCVPA study methods are reported elsewhere (24). In brief, participants included healthy infants 4-8 weeks old prior to first dose of PCV13, healthy children 18 weeks– 7 years old who received PCV13 as part of routine immunisation or the catch-up campaign, and healthy children 3–10 years old who were age-ineligible (born on or before 11 November 2010 and therefore too old) to receive PCV13. Epidemiological information collected include household location (GPS coordinates) and household composition, date of nasopharyngeal (NP) swab collection, participant’s age and gender, vaccination status, and socioeconomic status. Furthermore, we used population density of Blantyre from each year of the study obtained from WorldPop research programme(55). These population density data are modelled outputs from a statistical model that takes as inputs the national census data, geographical and settlement data to infer the locations where people live. The model predicts the number of people within a100m X 100m grid with associated confidence intervals

### Isolates and whole genome sequences

*S. pneumoniae* was isolated by culture from NP samples and bacterial DNA extracted from individual colonies as previously described (56). DraG assemblies of whole genome sequences were obtained from the PCVPA study and linked with study data from PCV-vaccinated children aged 2 to 7 years (n = 1,882) and PCV-unvaccinated (age ineligible) children aged 5 to 10 years (n = 600) collected in Blantyre from 2015 to 2019 (24).

### Gene4c typing and an4bio4c resistance tes4ng

Whole Genome sequences of pneumococcal isolates, serotypes, genetic lineages, and antimicrobial resistance were determined using Pathogenwatch (<u>hUps://pathogen.watch/</u>)(57). Lineages were defined by both the Global Pneumococcal Sequence Cluster (GPSC) from POPpunk v2.7.0 and the pneumococcal multilocus sequence type (MLST) scheme. The penicillin MIC was predicted *in silico* using the SPN-PBP-AMR machine learning algorithm using the EUCAST 2024 breakpoints (58–60). Other AMR gene and mutations were detected using Pathogenwatch AMR prediction module.

### Extrac4ng divergence 4me from Bayesian 4me calibrated phylogene4c tree

A Bayesian Tme-calibrated phylogenetic tree was constructed for each GPSC. To find the best reference sequence to align for each GPSC, we utilised ReferenceSeeker to find the closest related complete genome sequence from the NCBI Genbank database for each GPSC (61,62). These isolates were then aligned using Parsnp v1.0, and recombination events were removed with Gubbins v3.3.1 (63,64). A Bayesian Tme-calibrated phylogenetic tree was then constructed using BactDating v1.1.2 with the mixedcarc model (65). The tree was used for further analysis if all parameters converged and reached an effective sample size > 100.

Divergence distances between isolate pairs were extracted using the rrspread R package (hUps://github.com/hsuehchien66/rrspread_v2).

### Sta4s4cal analysis of pairwise distance and divergence from phylogene4c tree

A generalized additive mixed model (GAMM) with a gaussian distribution was constructed to analyse the pairwise divergence distances of isolates with less than 10 years of divergence. The model formula was specified as follows:

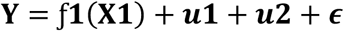

Where Y represents the pairwise distance (in km) between samples, ƒ1(X1) is a smooth function of the fixed effect for time divergence (in years) between pairs, is a random intercept accounting for the non-independence of the sample pairs, and ɛ denotes the residual error.

This model was constructed using the *mgcv* v1.9-1 R package.

The saturation point of each GAMM curve was defined by the first instance of the derivative of the curve being less than 0.1, indicating a flattening.

To calculate the relative risk (RR) of transmission across different divergence times and distances, these were binned into the following intervals: 0–1 year, 1–2 years, 2–3 years, 3–4 years, and 4–5 years; and 0-0.075km, 0.075-0.5km, 0.5-1km, 1-2km, 2-3km, 3-4km and 4-5km.

We implemented the following formula adapted from Cheng et al., 2024 (17):

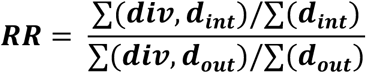

Where ∑(𝒅𝒊𝒗, 𝒅_𝒊𝒏𝒕_)is the sum of the number of pairs that fall within a specified divergence time interval and specified pairwise distance.,)∑(𝒅_𝒊𝒏𝒕_)is the sum of number of pairs within that specified distance interval, ∑(𝒅𝒊𝒗, 𝒅_𝒐𝒖𝒕_) is the sum of number of pairs with divergence times outside the specified distance and ∑(𝒅_𝒐𝒖𝒕_) is the sum of the number of pairs outside that specified distance interval.

The RR was also calculated based on the likelihood of transmission between children as a function of their age difference, grouped into the following intervals: 0-1, 1-2, 2-3, 3-4, 4-5- and 5-6-years difference. This RR was calculated using the following formula:

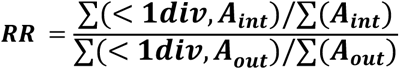

Where ∑(< 1 𝑑𝑖𝑣, 𝐴_&’(_) is sum of number of pairs<1 year divergence within a specified age difference, ∑(𝑨_𝒊𝒏𝒕_) is sum of number of pairs within a specified age difference, ∑(< 𝟏𝒅𝒊𝒗, 𝑨_𝒐𝒖𝒕_) the sum of number of pairs <1 year divergence outside a specified age difference and ∑(𝑨_𝒐𝒖𝒕_) is sum of number of pairs outside a specified age difference.

The confidence interval (CI) for the RR was estimated by bootstrapping 20 initiation, For each initiation we resampling the individuals with replacement and recalculating the RR for each resampled dataset.

A univariate and multivariable mixed-effects logistic regression model was developed to examine the significance of child epidemiological data, such as age of child household population data, social economic score, and population density as well as bacterial serotype and antimicrobial resistance (AMR) genotype variables, as predictors. The outcome variable was binary, coded as 1 for isolates that were found in at least one pair with divergence in time and distance below the saturation points identified from the GAMM model, indicating recent transmission events, and 0 otherwise. This was used to distinguish recent transmission of lineages in the study. The GPSC lineages were set as a random effect.

A univariate and multivariable logistic regression model were also constructed to explore the significance of human factors associated with carriage and non-carriage of pneumococcal. All continuous variables included in the models were standardised (z-scored) to normalise their scales. For both models, variables used for prediction were selected based on the absence of multicollinearity and a lower AIC score. Results were considered significant if P value < 0.05. mixed-effect logistic regression mode was constructed using the lme4 v1.1-35.4 R package.

### Random forest model

A random forest classifier model was constructed to classify a transmission event, and identify human and bacterial characteristics which predict transmission (20,66,67). Human characteristics included the age of the child, year of sample collected, socioeconomic score, household density (including number of children 5 years or younger, number of children 5-15 years or younger, and number of adults). Bacterial features included isolate’s GPSC, whether a PCV13 vaccine type serotypes, expected AMR phenotype from the genotype, and *in-silico* penicillin MIC. Categorical data were one-hot encoded. Optuna, a hyperparameter optimization framework (68), was used for hyperparameter optimisation, tuning parameters such as number of estimators, maximum depth, minimum samples split, minimum samples leaf, maximum features, class weight, and bootstrap settings, with performance evaluated via cross-validation using the mean ROC-AUC score. Feature selection was performed using Recursive Feature Elimination with Cross-Validation (RFECV) and 5-fold cross-validation. The best model was applied to training and testing datasets, with ROC-AUC, precision-recall AUC, and classification reports used to assess performance. The CI for performance metrics was calculated by bootstrapping 1,000 times. SHAP values were calculated on the train set and employed to identify important features, with partial dependence plots showing the impact of continuous variables. Python libraries Optuna v4.0, scikit-learn v1.5.2, and SHAP v0.46 were used.

### Lineage effect analysis

The lineage effect was analysed based on detected transmission within the population using the linear mixed model from pyseer v1.13.10 which use the method proposed by Earle et al (35,69). Genetic distance between isolates was calculated using Mash v2.0 and were assigned to their GPSC and MLST (70). Bonferroni correction was used to adjust p value for multiple comparison between different lineages. To determine the significant increase in prevalence of lineages we used the R stats package setting the denominator as all the isolates collected during those surveys and significance was determined by Chi-squared test for trend using rstatix package v0.7.2 R package.

### Detecting expansion effect population size

To determine clonal expansion and infer the effective population size over time, we used the Bayesian time-calibrated phylogenetic tree previously described and employed the CaveDive v0.1.1 R package, using priors from Helekal et al. (71).

## Data Availability

R and Python code used to plot the GAMM model and random forest model can be found in the Git repository: https://github.com/rorycave/Blantyre_SPN_geospace_paper. Whole genome sequence data are available from BioProject PRJNA1011974.

## Study ethical approval

The PCVPA study protocol received approval from the College of Medicine Research and Ethics Committee, University of Malawi (P.02/15/1677), and the Liverpool School of Tropical Medicine Research Ethics Committee (14.056). Written informed consent was obtained from adult participants and the parents or guardians of child participants. Children aged 8–10 years also provided informed assent. Consent included permission for publication.

## Author Summary

The pneumococcus is a leading bacterial cause of pneumonia, meningitis, and sepsis in children. Despite the widespread introduction of the pneumococcal conjugate vaccine in many lower- and middle-income countries, effective control of these diseases has not been achieved. Vaccine-targeted serotype carriage and disease continue to persist in these populations, accompanied by the emergence of antimicrobial-resistant lineages.

In this large, population-based study, we applied statistical and machine-learning approaches to integrate pneumococcal genomic, geospatial, and epidemiological data from Blantyre, Malawi. Our analysis identified key determinants of transmission including household proximity, child age, vaccine serotype, population density and penicillin susceptibility. Importantly, we found that it takes approximately four years for emerging lineages to became widespread across a population such as Blantyre, largely through transmission between neighbouring households. These findings support the need for enhanced vaccine strategies that target disease-causing and antimicrobial-resistant pneumococcal lineages, with a focus on pre- school children.

