## Supplemental Figures for "Characterising *Streptococcus pneumoniae* Transmission Patterns in Malawi Through Genomic and Statistical Modelling"

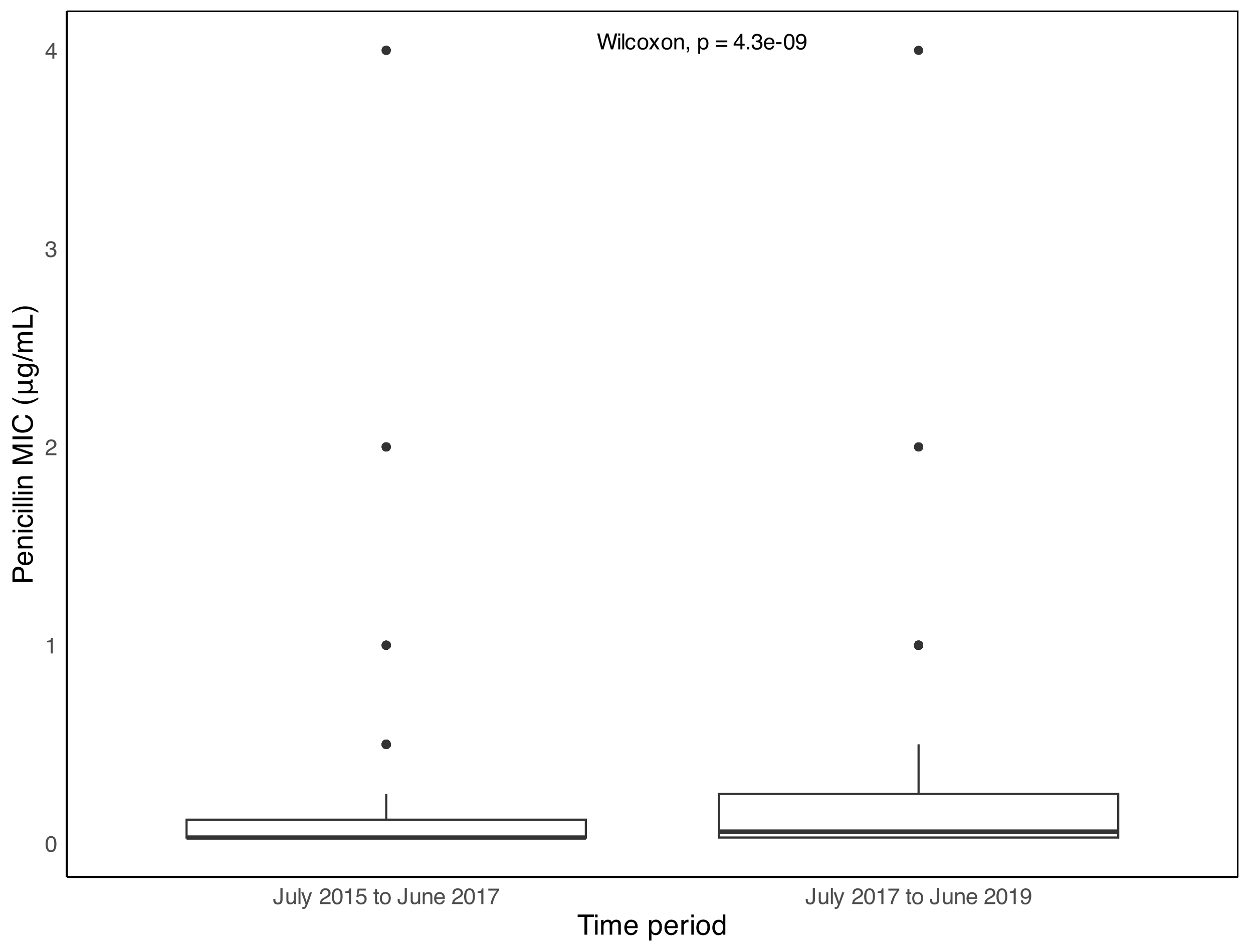

**Figure S1:** Boxplot showing Penicillin MIC change between early and late survey time points in the PCVPA dataset.

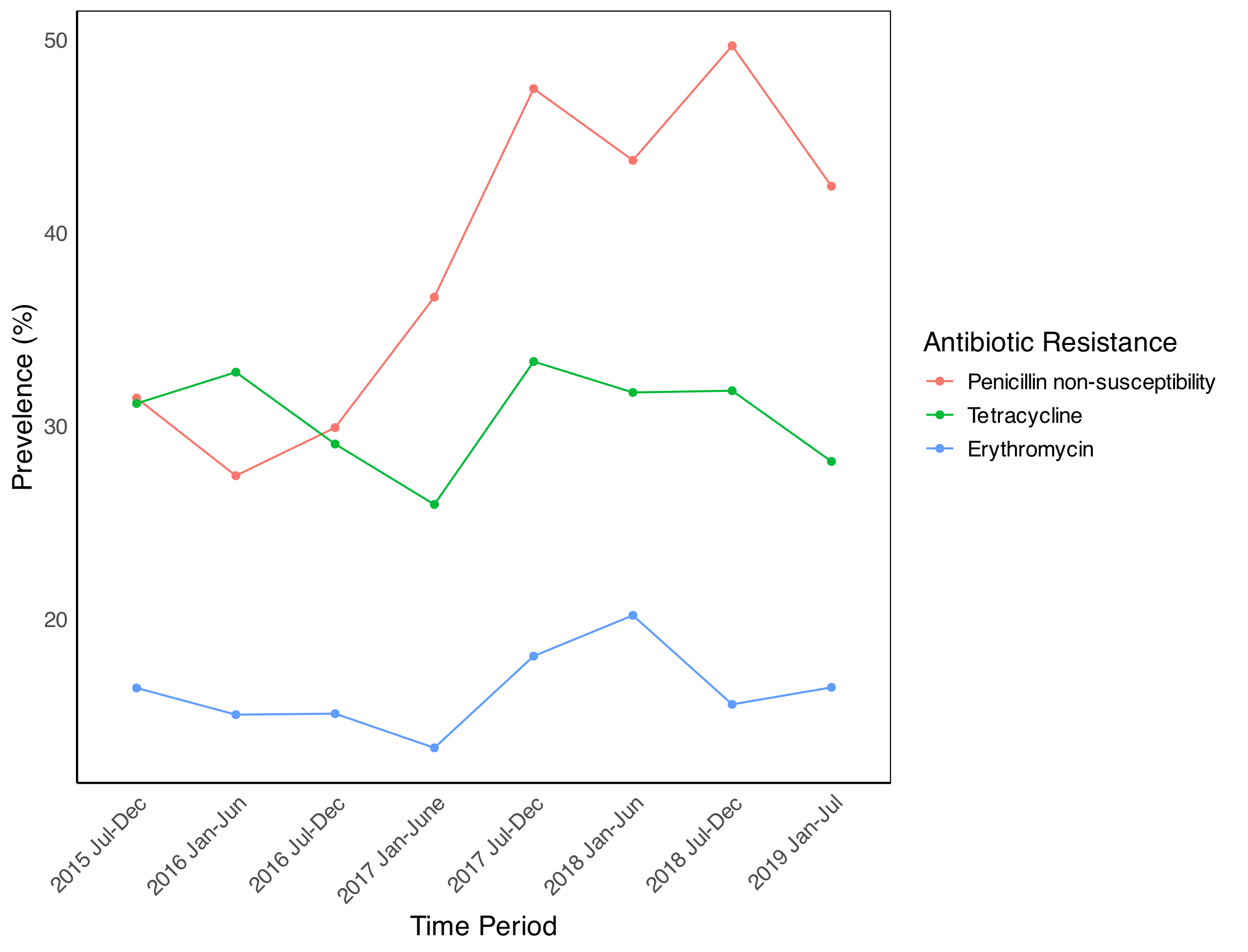

**Figure S2**: Line graph showing change of antibiotic resistance prevalence change during 8 separate time periods during the survey.

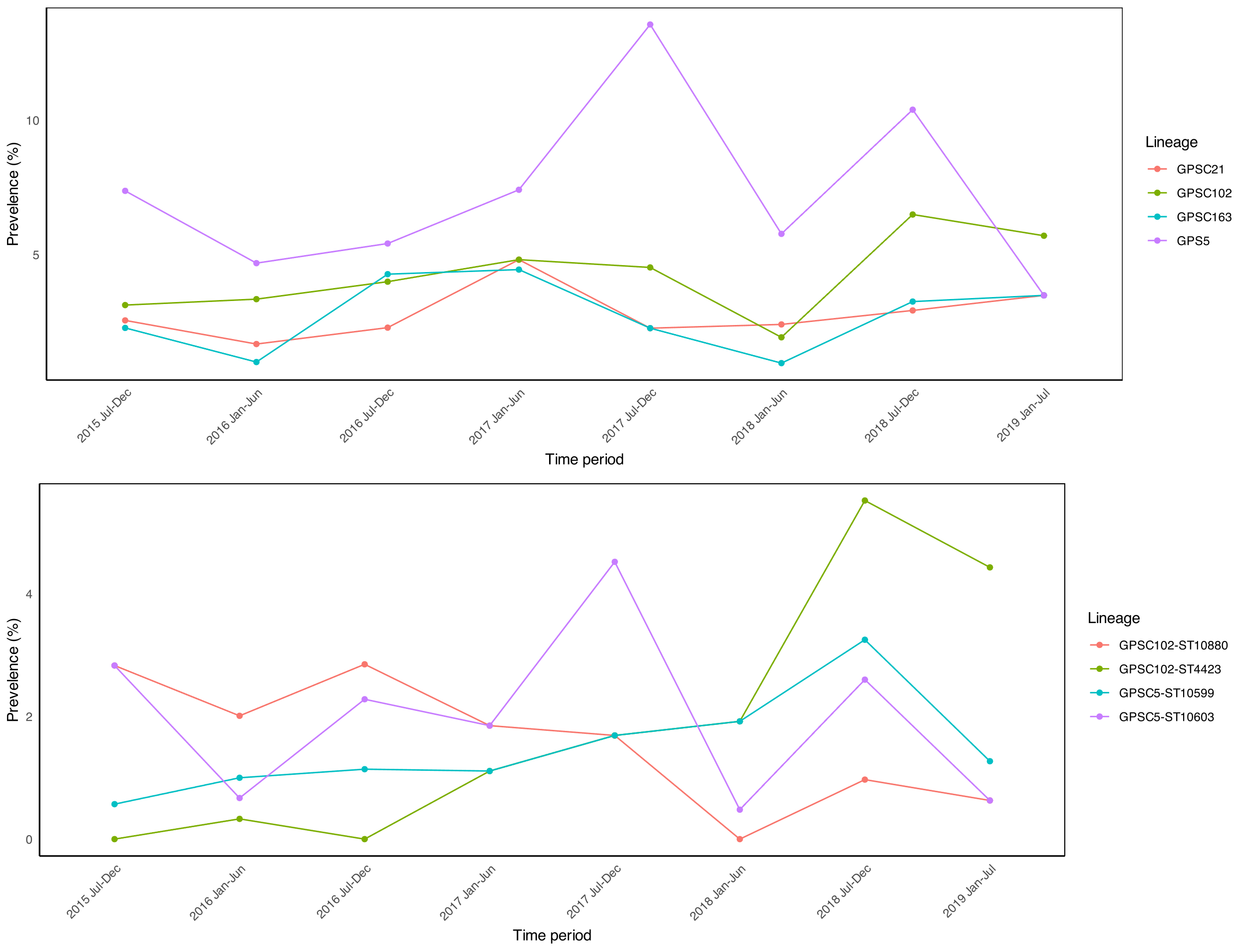

**Figure S3**: Line graph showing prevalence change of lineages that had significant lineage effect in transmission in the PCVPA dataset A) by their GPSC, B) By their MLST
